# Disproportionate impacts of COVID-19 in a large US city

**DOI:** 10.1101/2022.11.04.22281855

**Authors:** Spencer J. Fox, Emily Javan, Remy Pasco, Graham C. Gibson, Briana Betke, José-Luis Herrera Diestra, Spencer Woody, Kelly Pierce, Kaitlyn E. Johnson, Maureen Johnson-León, Michael Lachmann, Lauren Ancel Meyers

## Abstract

COVID-19 has disproportionately impacted individuals depending on where they live and work, and based on their race, ethnicity, and socioeconomic status. Studies have documented catastrophic disparities at critical points throughout the pandemic, but have not yet systematically tracked their severity through time. Using anonymized hospitalization data from March 11, 2020 to June 1, 2021, we estimate the time-varying burden of COVID-19 by age group and ZIP code in Austin, Texas. During this 15-month period, we estimate an overall 16.9% (95% CrI: 16.1-17.8%) infection rate and 34.1% (95% CrI: 32.4-35.8%) case reporting rate. Individuals over 65 were less likely to be infected than younger age groups (8.0% [95% CrI: 7.5-8.6%] vs 18.1% [95% CrI: 17.2-19.2%]), but more likely to be hospitalized (1,381 per 100,000 vs 319 per 100,000) and have their infections reported (51% [95% CrI: 48-55%] vs 33% [95% CrI: 31-35%]). Children under 18, who make up 20.3% of the local population, accounted for only 5.5% (95% CrI: 3.8-7.7%) of all infections between March 1 and May 1, 2020 compared with 20.4% (95% CrI: 17.3-23.9%) between December 1, 2020 and February 1, 2021. We compared ZIP codes ranking in the 75th percentile of vulnerability to those in the 25th percentile, and found that the more vulnerable communities had 2.5 (95% CrI: 2.0-3.0) times the infection rate and only 70% (95% CrI: 61%-82%) the reporting rate compared to the less vulnerable communities. Inequality persisted but declined significantly over the 15-month study period. For example, the ratio in infection rates between the more and less vulnerable communities declined from 12.3 (95% CrI: 8.8-17.1) to 4.0 (95% CrI: 3.0-5.3) to 2.7 (95% CrI: 2.0-3.6), from April to August to December of 2020, respectively. Our results suggest that public health efforts to mitigate COVID-19 disparities were only partially effective and that the CDC’s social vulnerability index may serve as a reliable predictor of risk on a local scale when surveillance data are limited.

## Introduction

The WHO estimates that the COVID-19 pandemic caused nearly 15 million excess deaths worldwide between its emergence in 2019 and the end of 2021. The burden fell disproportionately on countries in South-East Asia, Europe, and the Americas, with 68% of the estimated excess deaths occurring in 10 countries containing 35% of the global population [1]. In the United States, pandemic burden was initially concentrated around New York City, but spread geographically after the White House issued the *Opening Up America Again* guidelines in spring of 2020 [2]. The pandemic disproportionately harmed essential workers and racial and ethnic minority groups [3–6] as well as US counties [7–10] and cities [11–13] with high social vulnerability indices [14].

In response to these glaring disparities, scientists and public health leaders advocated for programs to support marginalized communities, including accessible testing facilities, community support programs to mitigate the socioeconomic, educational and healthcare harms resulting from lockdowns, proactive vaccination and antiviral campaigns, and effective public health communications [15–21]. Some but not all US vaccination campaigns successfully prioritized vulnerable geographic regions [22–29].

To prevent, detect, and reduce disparities in infectious disease burden, we need to increase the geographic and temporal resolution of our surveillance efforts, while reducing biases. Published estimates of COVID-19 burden in underserved populations are often derived directly from reported case or death counts, without correcting for ascertainment biases or disentangling risks of infection from risks of severe outcomes [7–9,30–40]. When available, both serological [41,42] and hospitalization data [43] can be used to estimate reporting rates. Several studies have highlighted the disproportionate burden of COVID-19 infections within cities [44,45], but only at single time points during the pandemic.

Here, we estimate the changing burden of COVID-19 at a local scale within a large US city throughout the first 15 months of the pandemic. Using ZIP-code and age-stratified hospitalization data, we track daily disparities in infection rates, hospitalization rates, and case reporting rates. As the SARS-CoV-2 virus continues to evolve along with our arsenal of medical and behavioral interventions, this method can help to ensure the reliability and equity of local risk assessments [46].

## Methods

We estimate the daily age-stratified numbers of infections for each of the 46 ZIP codes in Austin, Texas from hospital linelist data provided by the three major local healthcare systems to Austin Public Health [47]. As described below, we first estimate age-specific infection hospitalization rates (IHRs) from state-wide COVID-19 hospitalization data and SARS-CoV-2 seroprevalence data and then use the IHR estimates to infer time series of infections by age group and ZIP code using a deconvolution method similar to ref. [44].

### Estimating Texas statewide infection hospitalization rates (IHRs)

We analyzed age-stratified statewide COVID-19 hospitalization data [48] and statewide SARS-CoV-2 antibody seroprevalence data [49] to estimate the statewide infection hospitalization rate by age. Specifically, we estimate IHR for age group, ***k***, across the state of Texas (***μ***_***k***_,_***state***_) as:

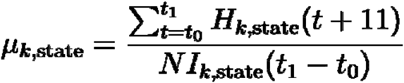

where ***t*** contains all dates between ***t***_**0**_ (July 29, 2020) and ***t***_**1**_ (May 27, 2021), ***H***_***k***,***state***_(*t* + 11) corresponds to all reported hospitalizations in age group ***k*** on day ***t* + 11** to account for the delay between infection and hospitalization [50,51], and ***NI***_***k***,***state***_ corresponds to the CDC’s estimated infections for Texas between the two dates [52] estimated as:

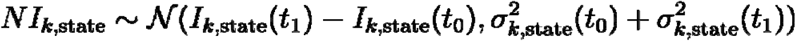

where 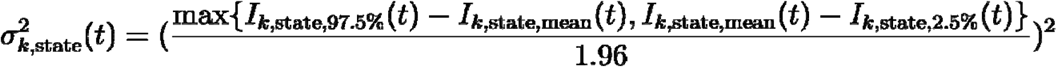 and corresponds to the variance of estimated infections at time ***t*** [52]. We aggregated hospital admission data, which are stratified into 0-17, 18-19, 20-29, 30-39, 40-49, 50-59, 60-69, 70-79, and 80+ year age groups, to match the stratification of the seroprevalence data (0-17, 18-49, 50-64, and 65+ years). For bins that do not align, we divided admissions evenly across years within a bin.

### ZIP- and age-specific infection hospitalization rates (IHRs)

Infection hospitalization rates depend on the underlying demographic makeup of a population [53]. To estimate age- and ZIP-specific IHRs from the statewide averages estimated above, we assumed that risk differences between ZIP codes could be captured by the proportion of the population estimated to be at high risk for severe COVID-19. We used a published methodology to estimate the proportion of the population at high risk for severe COVID-19 outcomes using data available from CDC’s PLACES for each ZIP code and the state on average [5,54–56]. We converted the statewide age-specific IHRs to ZIP-specific ones as:

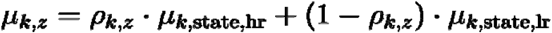

where ***μ***_***k***_,_***z***_ is the infection hospitalization rate for age group, ***k***, and ZIP code ***z, ρ***_***k***,***z***_ is the estimated age and ZIP code proportion of the population at high risk, and ***μ***_***k***_,_**state**,**hr**_ and ***μ***_***k***_,_**state**,**lr**_ are the statewide estimated age-specific IHRs for those at high and low risk to severe COVID-19 outcomes respectively. We assume a fixed hospitalization risk ratio between low and high risk individuals, ***μ***_***k***_,_**state**,**hr**_ = *η*_***k***_ · ***μ***_***k***_,_**state**,**hr**_, where ***η***_***k***_ is the age-specific hospitalization risk ratio estimated in [57]. For example, high risk individuals in the 20-24 and 75+ age groups are estimated to have 6.5 and 2.2 times the hospitalization risk respectively compared with low risk individuals in the same age group (Table S1). We then estimate ***μ***_***k***_,_**state**,**lr**_ and ***μ***_***k***_,_**state**,**hr**_ as:

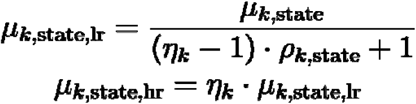

where ***μ***_***k***_,_**state**_ is the previously estimated statewide age-specific IHR, and ***ρ***_***k***_,_**state**_ is the statewide age-specific estimate for the proportion of the population at high risk for severe COVID-19. We then estimate ***μ***_***k***,***z***_ across Travis County using age- and ZIP-specific estimates for the proportion of the population at high risk for COVID-19. Confidence intervals for ***μ***_***k***,***z***_ are derived by propagating the estimated uncertainty from the age-specific statewide IHRs.

### Age- and ZIP code-specific infection estimates

The posterior distribution for infections (***I***_***k***,***z***_) in a specific age group (***k***) and ZIP code (***z***) was estimated as:

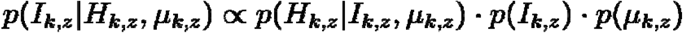

where ***<u>I</u>***_***<u>k,z</u>***_ indicates the total infection count, ***<u>H</u>***_***<u>k,z</u>***_ is the reported hospital admissions, and ***<u>μ</u>***_***<u>k,z</u>***_ is the age- and ZIP-specific IHR. Hospitalizations were assumed to be a binomial sample of the total infections governed by ***μ***_***k***,***z***_, ***μ***_***k***,***z***_ is assumed to follow a beta distribution, and ***I***_***k***,***z***_ follows a uniform discrete prior between the age- and ZIP-specific hospitalization count ***H***_***k***,***z***_ and population ***N***_***k***,***z***_:

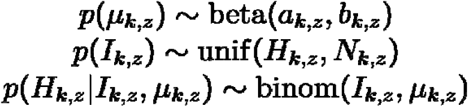

We estimate the parameters for the informative prior beta distribution, ***a***_***k***,***z***_ and ***b***_***k***,***z***_, using the ZIP and age-specific IHR estimates estimated from seroprevalence data in the previous section. Specifically we use the equation:

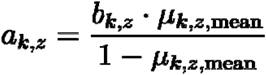

and identify the value of ***b***_***k***,***z***_ that minimizes the difference between ***μ***_***k***,***z*.2.5%**_ and the 2.5th percentile of the resulting beta distribution, **beta**(***a***_***k***,***z***,_ ***b***_***k***,***z***_). In essence, we estimate the shape parameters of a beta distribution that match the mean and lower bound estimate of the IHR. We used JAGS to sample 1,000 draws from ***p***(***I***_***k***,***z***_|***H***_***k***,***z***_,***μ***_***k***,***z***_) across four chains thinning every two samples and with a 200 sample burn-in period [58]. Throughout the paper we summarize the posterior distributions using their mean and 95% credible intervals.

We created a distribution of the time-series of infections from the 1,000 samples of ***p***(***I***_***k***,***z***_|***H***_***k***,***z***_,***μ***_***k***,***z***_) through the hospital admission timing and the delay distribution between infection and hospital admissions. Specifically, we fit a gamma distribution to the combined distribution derived from the time to symptom onset estimated in [50] and the time between symptom onset and hospital admission estimated in [51]. The combined distribution is estimated as **Г(shape= 2.99, rate = 0.27)**. For each infection in ***I***_***k***,***z***_, we draw a hospitalization timing from ***H***_***k***,***z***_**(*t*)**, the number of hospitalizations in an age- and ZIP code group on day ***t*** and assign an infection time from the estimated gamma distribution similar to the deconvolution method used in [44] without necessitating a smoothing function for the infection time-series. For age and ZIP code groups that had estimated infections but zero reported hospitalizations, we drew the infection timing from the full hospitalization time-series, ***H*(*t*)**, where ***H*(*t*) =∑ (*H***_***k***,***z***_**(*t*))** for day, ***t***, summed across all age groups and ZIP codes.

### Reporting rate estimates

We assumed reported cases are distributed binomially as given by

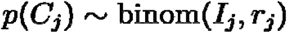

where ***j*** describes the specific subgroup of interest (age group, ***k***, and/or ZIP code, ***z***) for a specified time period, ***I***_***j***_ is the estimated infections, and ***ρ***_***j***_ is the estimated reporting rate. Assuming a uniform beta prior distribution on the reporting rate, the posterior for ***r***_***j***_ can be calculated as:

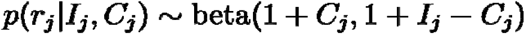

We estimated overall and subgroup reporting rates for the full study period through June 1, 2021 using cumulative age-specific case counts for Travis County [59] as well as ZIP code specific counts provided by Austin Public Health (APH) [60]. Separately, we estimated the age- and ZIP-specific reporting rates from a subset of testing data provided to us directly from Austin Public Health, which included 60% of reported cases in Travis County during the time period. Similar to the seroprevalence analysis, we lagged reported cases by 11 days to account for infection reporting delays [61,62].

### Infection estimate validation

To estimate how well our infection time-series approximate true infection rates in Travis county, we compare them to seroprevalence data for Texas estimated by the CDC [52] Regions in Texas faced different infection rates, so we adjusted Texas statewide seroprevalence estimates to Travis County as:

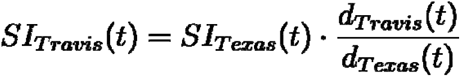

Where ***SI***_***Texas***_**(*t*)** is the seroprevalence infection estimate for Texas at time corrected for the delay in antibody positivity [52], ***d***_***Travis***_**(*t*)** and ***d***_***Texas***_**(*t*)** refer to the cumulative reported deaths in Travis County and Texas corrected by 20 days to account for the delay between infection onset and mortality [63].

### Social Vulnerability Index (SVI) as a predictor

The CDC’s Social Vulnerability Index (SVI) is a single indicator based on 15 different American Community Survey (ACS) variables that estimate a community’s ability to withstand environmental, biological and other stressors [14]. SVI values are given at the level of census tract as percentile ranks (range 0.0-1.0) within each state based on the 2014-2018 5-year ACS. For example, an SVI of 0.6 indicates that a census tract is more vulnerable than 60% of other census tracts in the state. Following [64], we aggregated SVI to ZIP codes using weighted averages based on the percent of residential addresses in a ZIP code that fall in each census tract [65].

We estimated the impact of SVI on infection and reporting rates using a mixed effect poisson regression model using the lme4 R package [66]. For estimating the impact of SVI on infection rates the model can be described as:

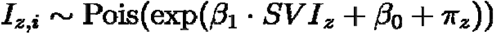

Where ***I***_***z***,***i***_ is the infection ***ith*** infection estimate sample estimated for ZIP code, ***z, SVI***_***z***_ is the ZIP codes’ SVI, ***π***_***z***_ is the ZIP code level random effect, ***β***_**1**_ is the fixed effect of SVI on infections, and ***β***_**0**_ is an intercept term. We use the ZIP code population as an offset in the model to standardize infection rates. For estimating the impact of SVI on reporting rates the model can be described as:

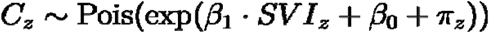

where ***C***_***z***_ is the reported case count for ZIP code, ***z, SVI***_***z***_, is the ZIP codes’s SVI, ***π***_***z***_ is the ZIP code level random effect, ***β***_**1**_ is the fixed effect of SVI on cases, and ***β***_**1**_ is an intercept. We use the 1,000 ZIP code infection estimate samples as an offset in the model to standardize reporting rates. For the age and ZIP code-stratified analysis we also include an interaction term between age and SVI, so the equations become:

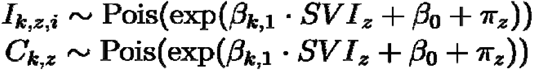

where ***I***_***k***,***z***,***i***_ is the ***ith*** infection estimate sample in age group, ***k*** for ZIP code, ***z, C***_***k***,***z***_, is the reported cases in age group, ***k*** for ZIP code, ***z***, and ***β***_***k***,**1**_ is the SVI regression coefficient for age group, ***k***. For the infection and reporting rate models we use the age- and ZIP-population and infection estimates as an offset respectively.

The SVI regression coefficients, ***β***_**1**_ and ***β***_***k***,**1**_, can be interpreted as inequality metrics, quantifying the relative infection and reporting risks as a function of SVI. Because there are no ZIP codes with a value of 0 or 1 for SVI in our sample, we report the relative infection and reporting rates between ZIP codes in the 25th and 75th percentile in Travis County throughout the manuscript.

## Results

We analyzed spatial COVID-19 burdens in Austin, Texas using hospital admission data from March 11, 2020 to June 1, 2021. This period preceded the emergence of the Delta variant and included a small wave in April 2020, followed by larger waves in the summer and winter (Figure 1A). As of June 1, 2021, there were 83,722 reported cases, 6,474 hospitalized patients, and 1,024 deaths of COVID-19 in Travis County, which has 1.3 million residents, covering 57% of the Austin metropolitan area population. We estimate that 16.9% (95% CrI: 16.1-17.8%) of the population were infected in this time period and 34.1% (95% CrI: 32.4-35.8%) of all infections were reported. These estimates are consistent with the CDC’s official seroprevalence estimates (Figure 1B).

**Figure 1:**
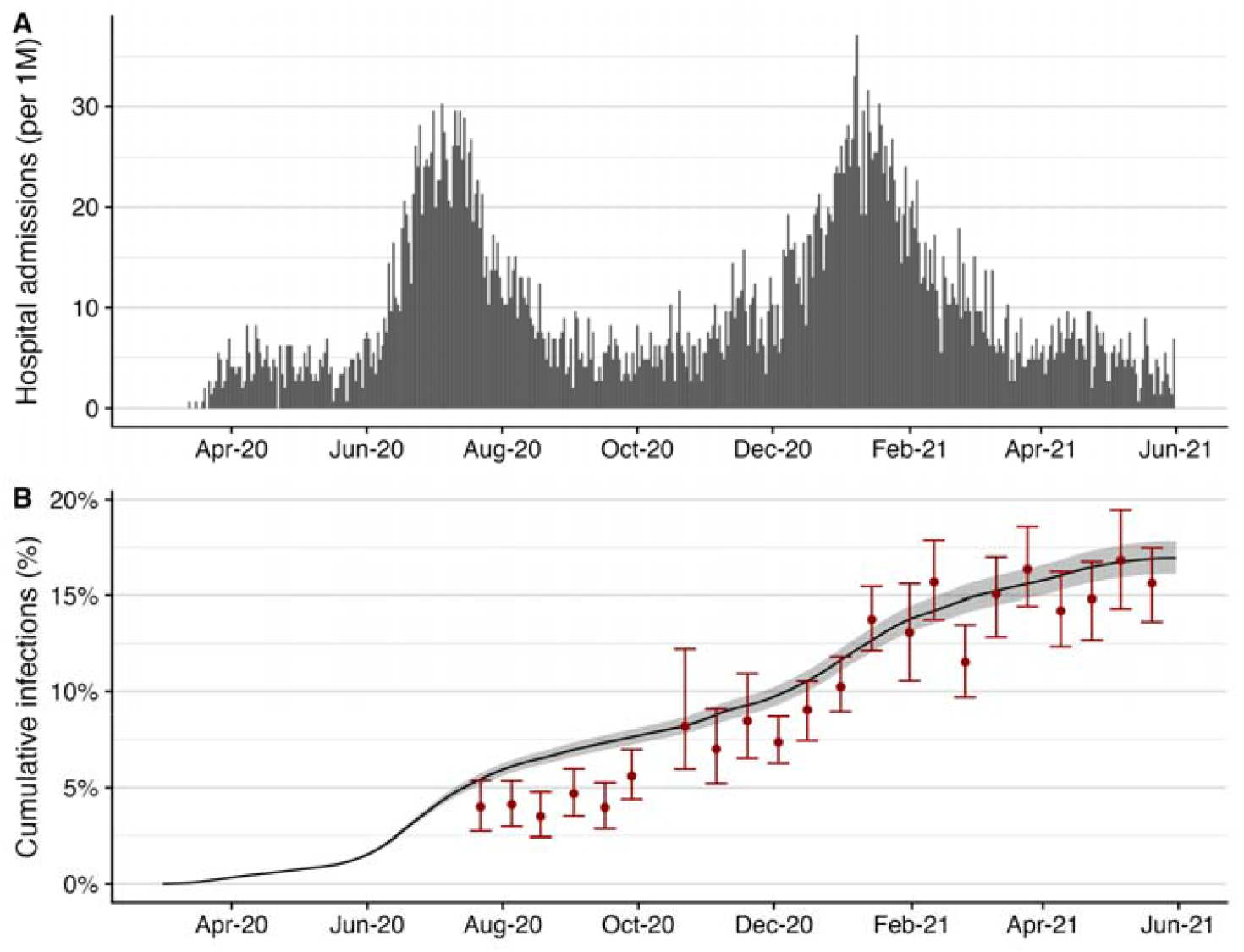
COVID-19 hospital admissions and estimated cumulative infections for Travis County (Austin, TX) from March 1, 2020 to June 1, 2021. (A) Daily reported COVID-19 hospital admissions per 1 million residents [68]. (B) Estimated cumulative infections with 95% credible intervals (black line and gray ribbon) compared to prior seroprevalence-based estimates (red points and error bars) [69].

Statewide, we estimate that one in 435 (95% CI: 244-625) infections in individuals aged 0-17 years and one in 4.9 (95% CI: 3.0-6.8) infections in individuals over age 65 led to hospitalization (Table 1). This is consistent with published estimates from China [67] and France [43] (Figure S2). In Travis County, children aged 0-17 experienced the lowest hospitalization rate, with 50.3 hospital admissions per 100,000, and adults over age 65 experienced the highest hospitalization rate of 1,381 per 100,000 (Figure 2A). In contrast, reported cases were relatively similar across age groups, ranging from 3,793 per 100,000 in children to 7,159 per 100,000 in young adults (Figure 2B).

**Table 1:** SARS-CoV-2 infection hospitalization rate (IHR) across Texas estimated from statewide seroprevalence and hospitalization data from July 29, 2020 through May 27, 2021.

| Age group | Number of COVID-19 hospital admissions [48] | Estimated infections based on seroprevalence data (95% CI) [49,52] | Estimated IHR (95% CI) |
| --- | --- | --- | --- |
| 0-17 | 4,868 | 2,076,780 (1,183,462-2,970,098) | 0.23% (0.16%-0.41%) |
| 18-49 | 67,708 | 3,784,332 (2,842,976-4,725,687) | 1.79% (1.43%-2.38%) |
| 50-64 | 58,375 | 1,084,993 (708,561-1,461,425) | 5.38% (3.99%-8.24%) |
| 65+ | 100,671 | 495,141 (301,983-688,299) | 20.33% (14.63%-33.34%) |

**Figure 2.**
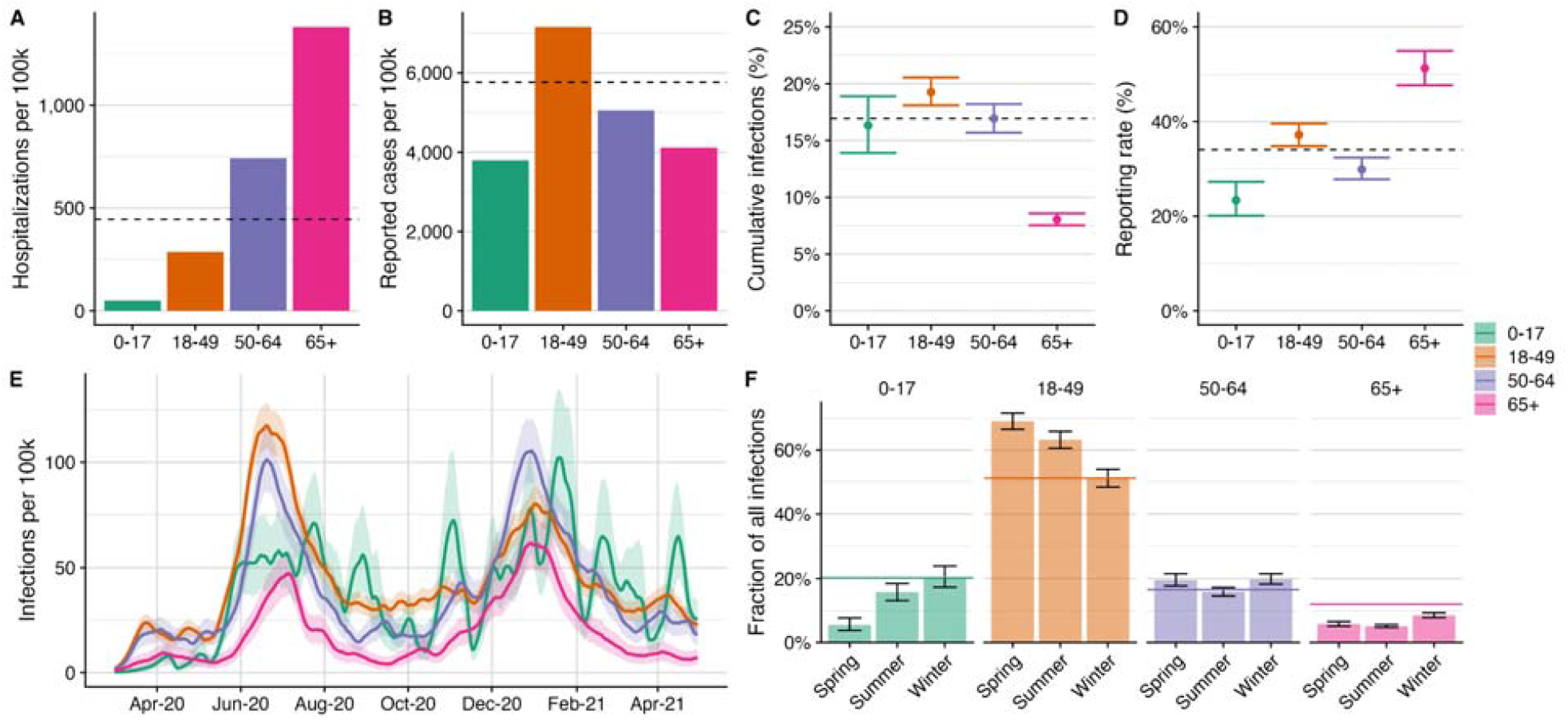
Estimated age-stratified COVID-19 burden in Travis country through June 1, 2021. (A) Reported COVID-19 hospital admissions by age group. (B) Reported COVID-19 cases by age group. (C) Estimated percent infected by age group. (D) Estimated COVID-19 case reporting rates by age group up to June 1, 2021. In (A)-(D), horizontal dashed lines indicate county-wide average rates. (E) Estimated daily infection rates (line) and 95% credible intervals (ribbons) by age group. (F) Distribution of infections across age groups for each period of the epidemic. The spring period refers to the two-month time period before the first major wave from March 1, 2020 to May 1, 2020, the summer period refers to the two-month period containing the first major wave from June 1, 2020 to August 1, 2020, and the winter period refers to the two-month period containing the second major wave from December 1, 2020 until February 1, 2021. Bars indicate the fraction of all infections during the time period in each Age group, with the error bars indicating the 95% credible intervals. The horizontal colored lines in panel F indicate the proportion of the Travis county population in the specified age group.

By June 1, 2021, we estimate that 19.3% (95% CrI: 18.1-20.6%) of 18-49 year olds were infected, while only 8.0% (95% CrI: 7.5-8.6%) of individuals over age 65 were infected (Figure 2C). The estimated percent of cases reported increases with age, ranging from 23.4% (95% CrI: 20.1-27.3%) in 0-17 year olds to 51.3% (95% CrI: 47.7-54.9%) in over 65 year olds (Figure 2D).

All age groups experienced two large waves during the study period, though the summer 2020 was relatively mild for children (Figure 2E). Relative infection rates across age groups evened out over time (Figure 2F). For example, children, who account for 20.3% of the Travis county population, constituted 5.5% (95% CrI: 3.8-7.7%) of all infections between March 1, 2020 and May 1, 2020 and 20.4% (95% CrI: 17.3-23.9%) of all infections between December 1, 2020 and February 1, 2021. The proportion of infections occurring in 18-49 year olds, who make up 51.2% of the population, dropped from 69.1% (95% CrI: 66.5-71.6%) during the spring 2020 period to 51.3% (95% CrI: 48.4-54.0%) during the winter 2020-2021 wave (Figure 2F and S3). Reported case and hospitalization counts do not clearly exhibit this reversal in age-specific risks (Figure S4 and S5).

Estimated COVID-19 burden varies significantly across ZIP codes within Travis County, with Interstate 35 roughly partitioning the county into high risk ZIP codes in the East and low risk ZIP codes in the West (Figure 3A and 3B). High COVID-19 risk visibly aligns with high social vulnerability, as measured by ZIP-code level SVI (Figure 3C). Our estimates for ZIP-code level infection hospitalization rates exhibit the opposite geographic trend (Figure 3D) from the absolute hospitalization rates (Figure S6). We estimate that downtown Austin (78701) had the highest infection rate and lowest reporting rate of any ZIP code, with an estimated 39.2% (95% CrI: 29.6-50.2%) of the ZIP code infected and only 18% (95% CrI: 14-23%) of infections reported (Figure 3E-3F). In contrast, a Southwest Austin ZIP code (78739) had the lowest estimated infection rate of 4.8% (95% CrI: 2.6-8.5%), and a West Austin ZIP code (78732) had the highest reporting rate of 69% (95% CrI: 39-97%). Similar geographic patterns exist for each of the four age groups (Figure S7-S8).

**Figure 3.**
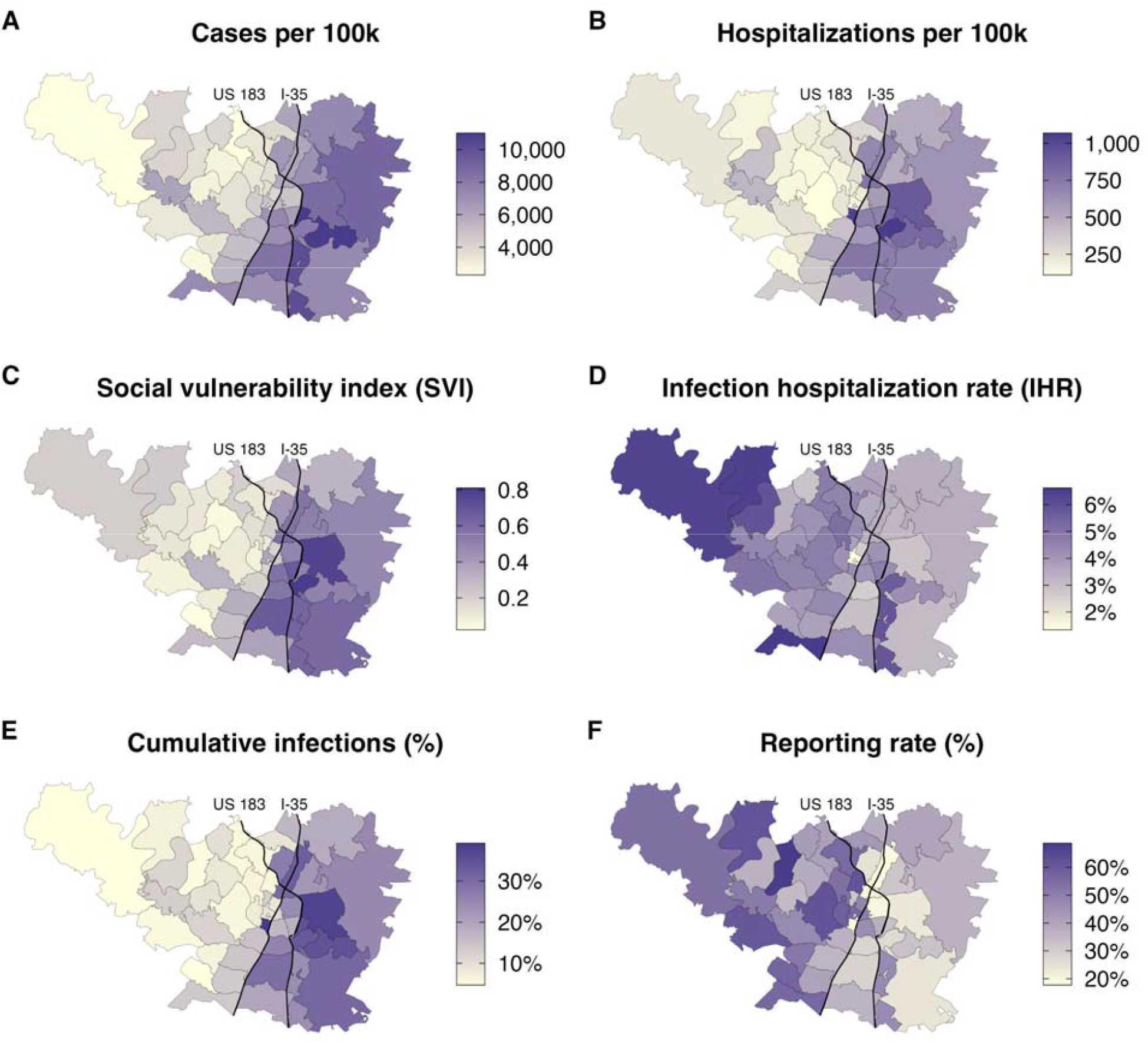
Reported and estimated COVID-19 burden by ZIP code for Travis County between March 1, 2020 and June 1, 2021. (A) Reported COVID-19 cases per 100,000. (B) Reported COVID-19 hospitalizations per 100,000. (C) Social Vulnerability Index [14] (D) Estimated infection hospitalization rate (IHR). (E) Estimated cumulative infections as of. (F) Estimated percent of COVID-19 infections that were reported. Thin black curves indicate Interstate 35 and highway US 183.

The cumulative infection rates, case rates, and hospitalization rates are positively correlated with social vulnerability across Travis County’s 46 ZIP codes (Figure 4A, Figure S9). We compare the relative risks for individuals living in a ZIP code at Travis County’s 25th (SVI= 0.12) and 75th (SVI = 0.5) percentile by SVI, where higher SVI indicates higher social vulnerability. Controlling for random ZIP code-level effects, we estimate that ZIP codes in the 75th SVI percentile experienced 2.5 (95% CrI: 2.0-3.0) times the infection rate of those in the 25th percentile. Similar trends are observed for each age group, with all relationships estimated to be statistically significant (Figure S10, Table S2). COVID-19 burden is often estimated directly from reported case or hospitalization data, without correcting for geographic biases in testing and underlying risk factors. For Travis county, we find that the subset of case data from APH provides a reasonable approximation but hospitalization data tends to inflate the estimated disparities (Table S2). We aggregate the estimated number of infections occurring in each ZIP code into four-week periods from March 1, 2020 to June 1, 2021, and measure the relationship between SVI and the relative infection risk during this period. Significant disparity (i.e., a relative risk greater than one) persisted throughout the period and was highest during the first three months of the pandemic (Figure 4B). In April 2020, individuals living in the 75th SVI percentile ZIP code had an expected 12.3 (95% CrI: 8.8-17.1) times greater infection risk than those living in the 25th percentile SVI ZIP code. This ratio declined to 4.0 (95% CrI: 3.0-5.3) in August 2020 and to 2.7 (95% CrI: 2.0-3.6) in December 2020.

**Figure 4.**
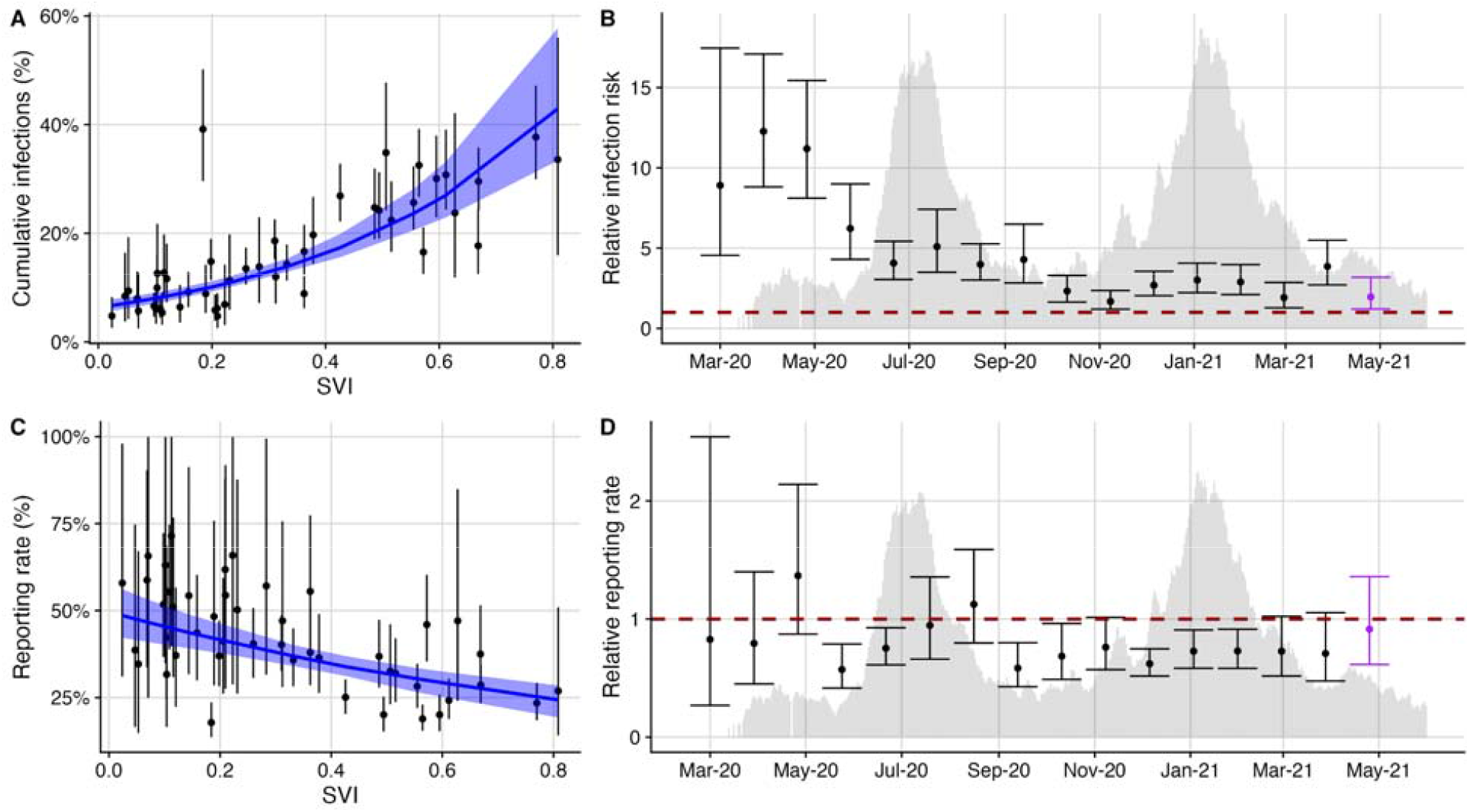
Infection and reporting rates correlate with social vulnerability in Travis County from March 1, 2020 to June 1, 2021. (A) Across the 46 ZIP codes, SVI is a significant predictor of estimated cumulative infections (p<0.001). The blue line and ribbon indicate the mean and 95% prediction interval from the fitted Poisson mixed-effects model. (B) Using the fitted model, we compare the expected infection rates among more and less vulnerable ZIP codes (specifically, ZIP codes at the 75th and 25th percentiles in the SVI distribution, respectively). The points indicate the expected ratio between these two values calculated using the estimated SVI regression coefficient from the 4-week time period; error bars indicate 95% CI’s. (C) Across the 46 ZIP codes, SVI is a significant predictor of estimated case reporting rates (p<0.001). The blue line and ribbon indicate the mean and 95% prediction interval from the fitted Poisson mixed-effects model. (D) Four week estimate for the inequality relationship between SVI and infection reporting rates across the 46 ZIP codes. Points and error bars show the mean and 95% CI for the relative reporting rate of individuals living in ZIP codes in the 75th SVI percentile compared with those living in the 25th SVI percentile. The red, horizontal dashed lines in B and D indicate if there were equitable infection risks or reporting rates across the 75th and 25th SVI percentile ZIP codes in the four week period. We overlay hospital admission time-series in B and D to showcase how inequality estimates compare with the progression of the local epidemic. For the final period (purple points and error bars), we removed the 10% of ZIP codes reporting zero infections to stabilize our regression estimates.

COVID-19 case reporting rates are negatively correlated with social vulnerability. We estimate that infections occurring in the 75th SVI percentile ZIP code were only 70% (95% CrI: 61%-82%) as likely to have been reported than those occurring in the 25th SVI percentile ZIP code. We further stratified by age group using a small sample of age-specific case data reported by the Austin Public Health community testing programs, which targeted vulnerable populations in East Austin (Figure S12) [70]. We found that the negative correlation between SVI and case reporting rates held for all age groups except those over 65 years, perhaps because of Austin’s efforts to improve testing access for high risk individuals (Figure S11, Table S2). Throughout the study period, the estimated ratio in reporting rates between the 75th and 25th SVI percentile ZIP codes fluctuated, often dropping to levels significantly less than one (Figure 4D).

## Discussion

In the US, the first wave of the COVID-19 pandemic disproportionately harmed essential workers [5,6], residents of long-term care facilities [71], racial and ethnic minority populations [72], and socially vulnerable neighborhoods within cities [31,44,73,74]. Public health agencies and government officials have tried to address these disparities through targeted testing, vaccination, distribution of personal protective equipment, information campaigns, and paid sick leave [15–19]. Using a new method for inferring infection risks and reporting rates from COVID-19 hospital admissions data, we demonstrate that disparities persist on a granular scale within a large US city throughout the first year of the pandemic.

Our estimates for the spatial burden of COVID-19 in Austin, Texas suggest that children were less likely to be infected than adults under age 65 during the first major wave of transmission in the summer of 2020 but not during the subsequent winter wave. This is consistent with prior estimates [53,75–77] and may be attributable to early school closures, strict compliance with social distancing measures [78], or the emergence of variants that more efficiently infect children [79,80]. We also find that individuals over age 65 generally had the lowest risks of infection, despite suffering the highest per capita hospitalization rate, which may stem from heightened precautionary behavior and other protective measures such as COVID-19 screening in long-term care facilities [81]. Our age-stratified estimates of cumulative SARS-CoV-2 incidence are generally consistent with those derived from seroprevalence data [49]. Reporting rates were lowest in children and highest in older adults (Figure 2), which may stem from the positive correlation between age and symptom severity [75].

Historically marginalized populations in the “Eastern Crescent” of Austin were disproportionately harmed throughout the first year of the pandemic [82,83], mirroring disparities reported for Santiago, Chile and New York City [44,45,73]. After controlling for the higher prevalence of underlying risk factors in more vulnerable communities, we find that the ZIP codes ranking in the 75th percentile of social vulnerability had a more than twofold higher infection rate and a roughly 70% the case reporting rate than those ranking in the 25th percentile. Our estimates for inequity in infection risk are significantly lower than those from raw hospitalization rates, which do not account for variation in the prevalence of comorbidities. The estimated ratio in infection risk between more and less vulnerable regions decreased significantly during the first four months of the pandemic, perhaps because of local efforts to increase access to SARS-CoV-2 testing, isolation facilities, critical health information, and eventually vaccines [84]. The apparent decrease in disparity may also stem from higher infection rates in vulnerable populations leading to a more rapid buildup of immunity or relatively higher infection rates in less vulnerable areas during later time periods [85,86]. As of June of 2021, however, there remained a significant gap in COVID-19 risks and burden which informed targeted efforts by Austin Public Health to increase access to tests, vaccines, information and COVID-19 healthcare.

We note several assumptions of analysis. First, the hospital admission data are limited by the accuracy of patient ZIP codes. Fewer than 1% of patients had unknown addresses. However, the missing data may correspond to vulnerable subgroups, such as people experiencing homelessness or undocumented residents, and thus obscure critical geographic or socioeconomic hotspots in our analysis. Second, we estimated each age- and ZIP-specific group independently rather than combining information across groups. This increases the uncertainty of our estimates but avoids the challenge of incorporating the changing contact and mobility patterns within the city throughout the pandemic [87–89]. Third, although we conducted analyses at a higher spatial resolution than most prior studies of COVID-19 burden, disparities in risk often occur at even more local scale [90]. Achieving COVID-19 health equity will require more granular surveillance and risk mitigation approaches. Finally, since we restricted our analysis to the first year of the pandemic, prior to the emergence of the Delta variant, we made the simplifying assumption that the infection hospitalization rates remained constant. Any estimates beyond this period would have to account for changes in severity resulting from the build up of vaccine-acquired and infection-acquired immunity and the evolution of the virus.

We estimate that less than 20% of the Austin, Texas population was infected by SARS-CoV-2 prior to June 1, 2021 and that vulnerable communities in East Austin bore the brunt of the first two large waves of transmission. Our study introduces a framework for tracking infection and reporting rates on a granular scale using hospitalization data and provides evidence that the CDC’s social vulnerability index (SVI) is a strong predictor of risk that can inform targeted interventions.

## Data Availability

All data are available upon reasonable request to Austin Public Health.

## Supplemental Information

**Table S1:** Relative hospitalization rates for high risk individuals compared with low risk individuals from [57].

| Age group | Relative hospitalization risk for high risk individuals |
| --- | --- |
| 0-0.5y | 6.03 |
| 0.5-4y | 6.03 |
| 5-9y | 6.03 |
| 10-14y | 6.48 |
| 15-19y | 6.48 |
| 20-24y | 6.48 |
| 25-29y | 6.48 |
| 30-34y | 5.5 |
| 35-39y | 5.5 |
| 40-44y | 4.625 |
| 45-49y | 4.625 |
| 50-54y | 3.78 |
| 55-59y | 3.78 |
| 60-64y | 3.24 |
| 65-69y | 3.24 |
| 70-74y | 2.32 |
| 75y+ | 2.19 |

**Figure S1:**
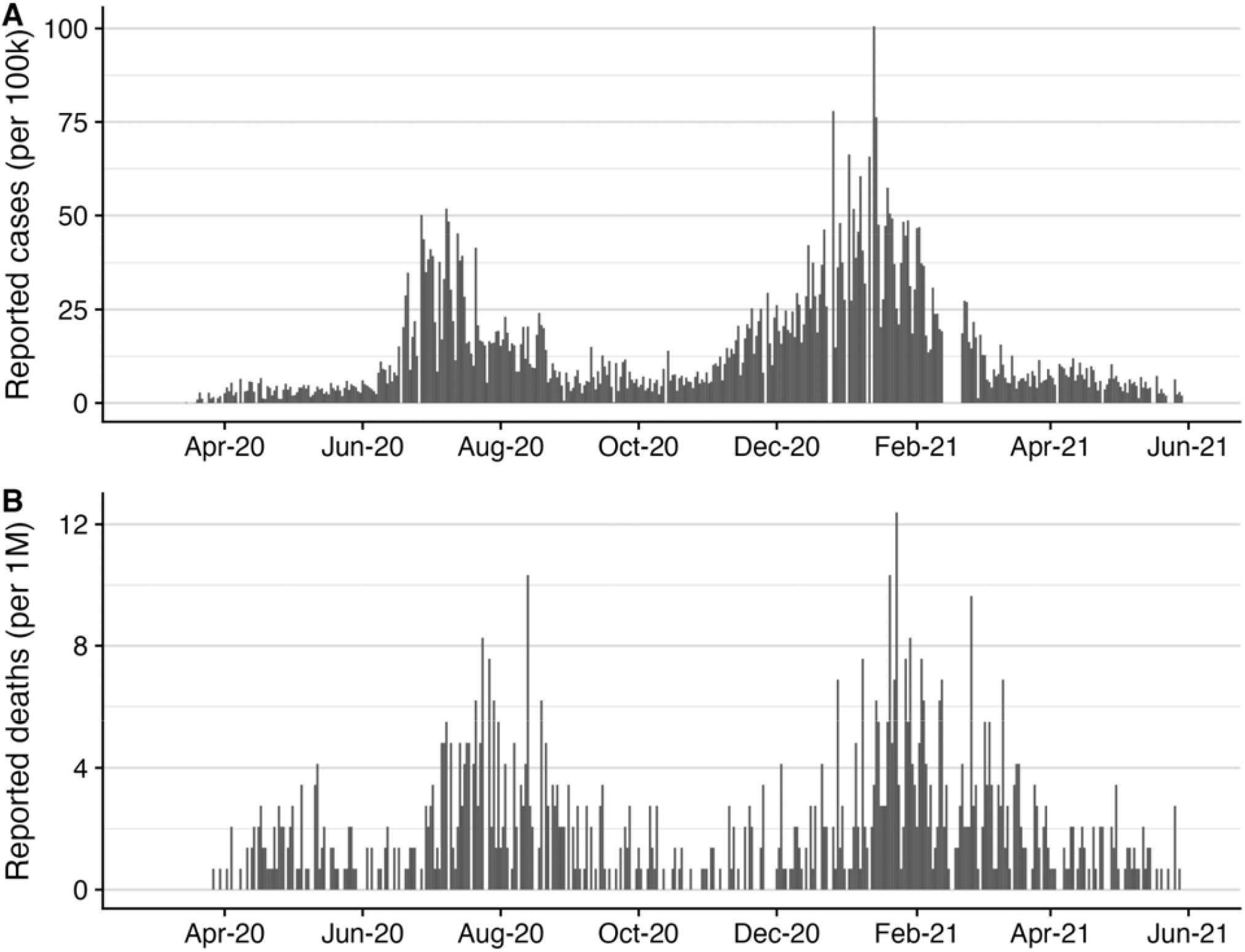
Daily COVID-19 burden estimates for Travis County, Texas from March 1, 2020 until June 1, 2021. Daily new reported case (A) and mortality (B) counts as reported by the New York Times for Travis County, Texas [59].

**Figure S2:**
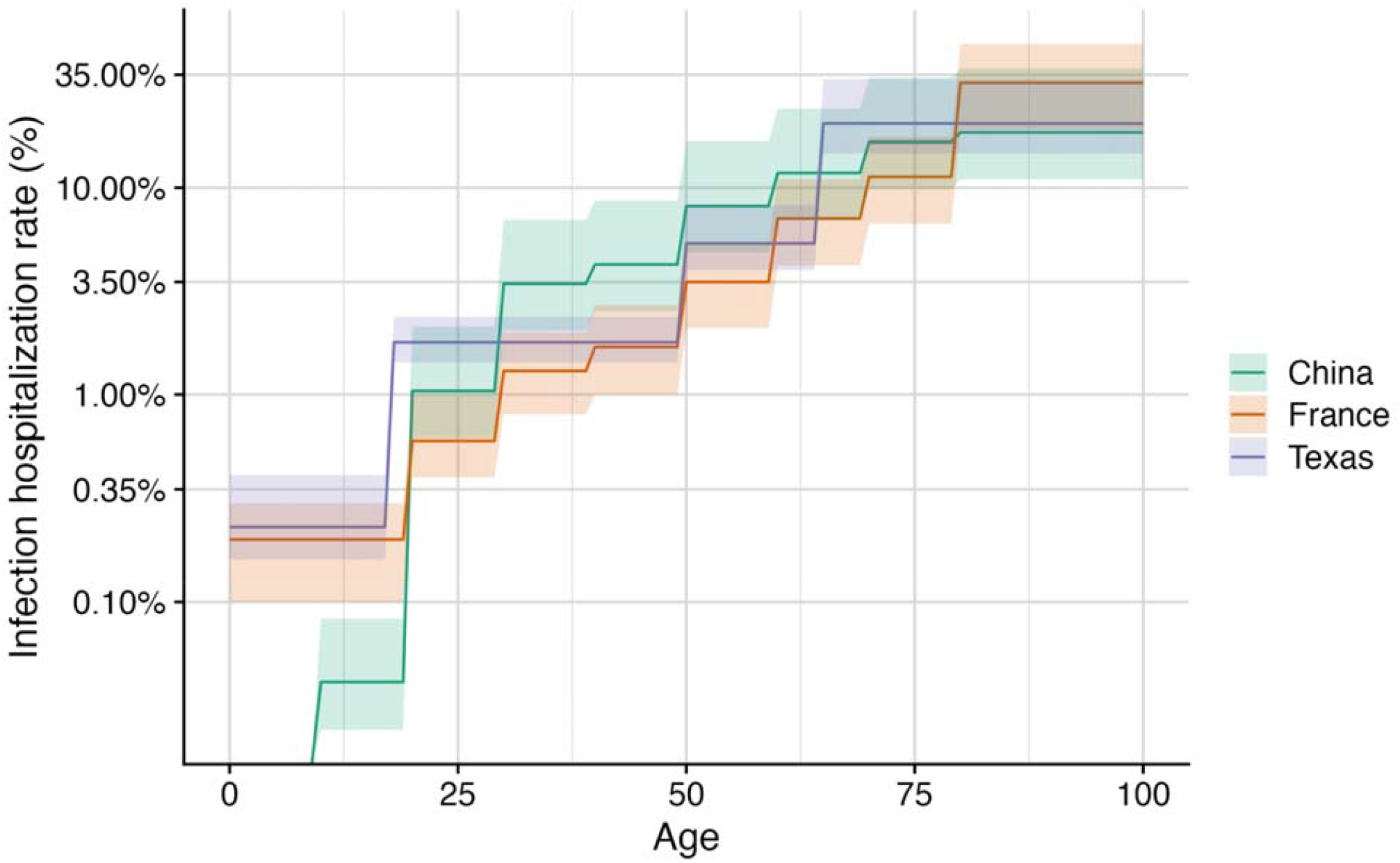
Comparison of age-dependent estimates for infection-hospitalization rates. Age-stratified estimates of the risk of severe COVID-19 (defined as risk for hospitalization) from China [67], France [43].

**Figure S3:**
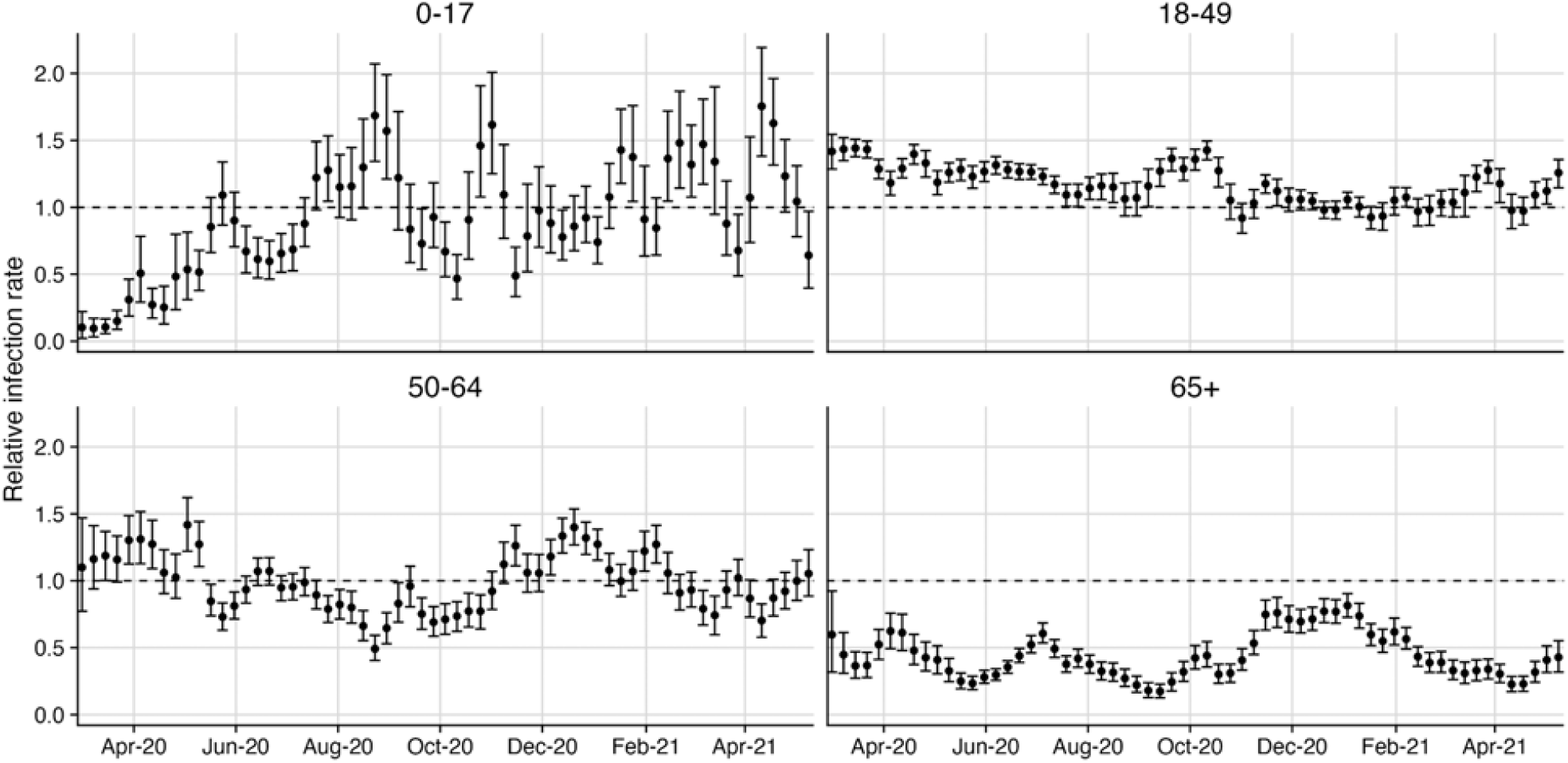
Weekly estimated relative infection rates from March 1, 2020 until June 1, 2021 across age groups. Points and error bars indicate the median and 95% confidence interval for the weekly infection rate with the size of the population. Values of 1 (horizontal dashed line) indicate that the fraction of the infections occurring that week equals the population fraction for the specific age group, while values below or above one indicate the age group faced disproportionately low or high infection risk respectively during that week. Only the 65+ age group consistently experienced disproportionately low infection rates compared with their population size over the whole pandemic.

**Figure S4:**
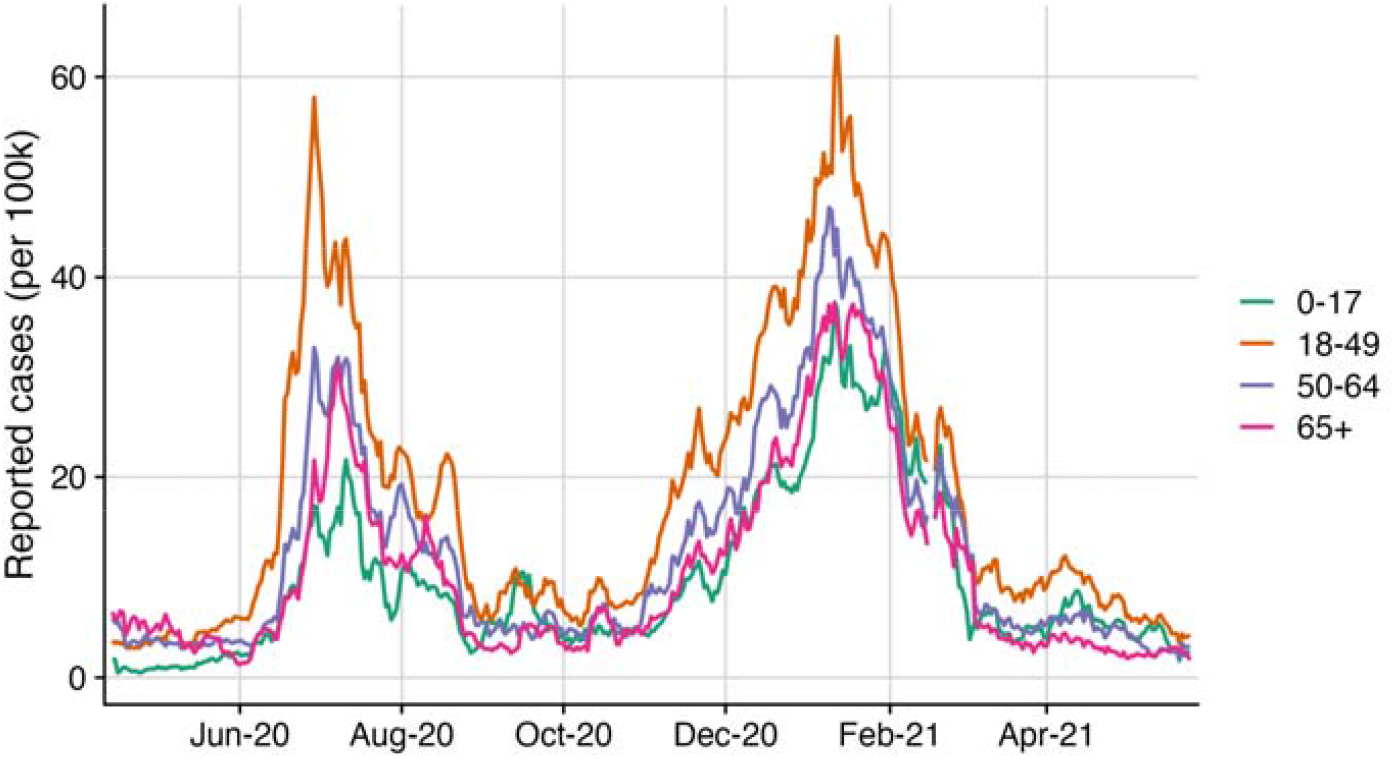
Reported 7-day average of case counts by age group from April 22, 2020 until May 28, 2021. Daily reported cases counts for each age group provided by Austin Public Health [60].

**Figure S5:**
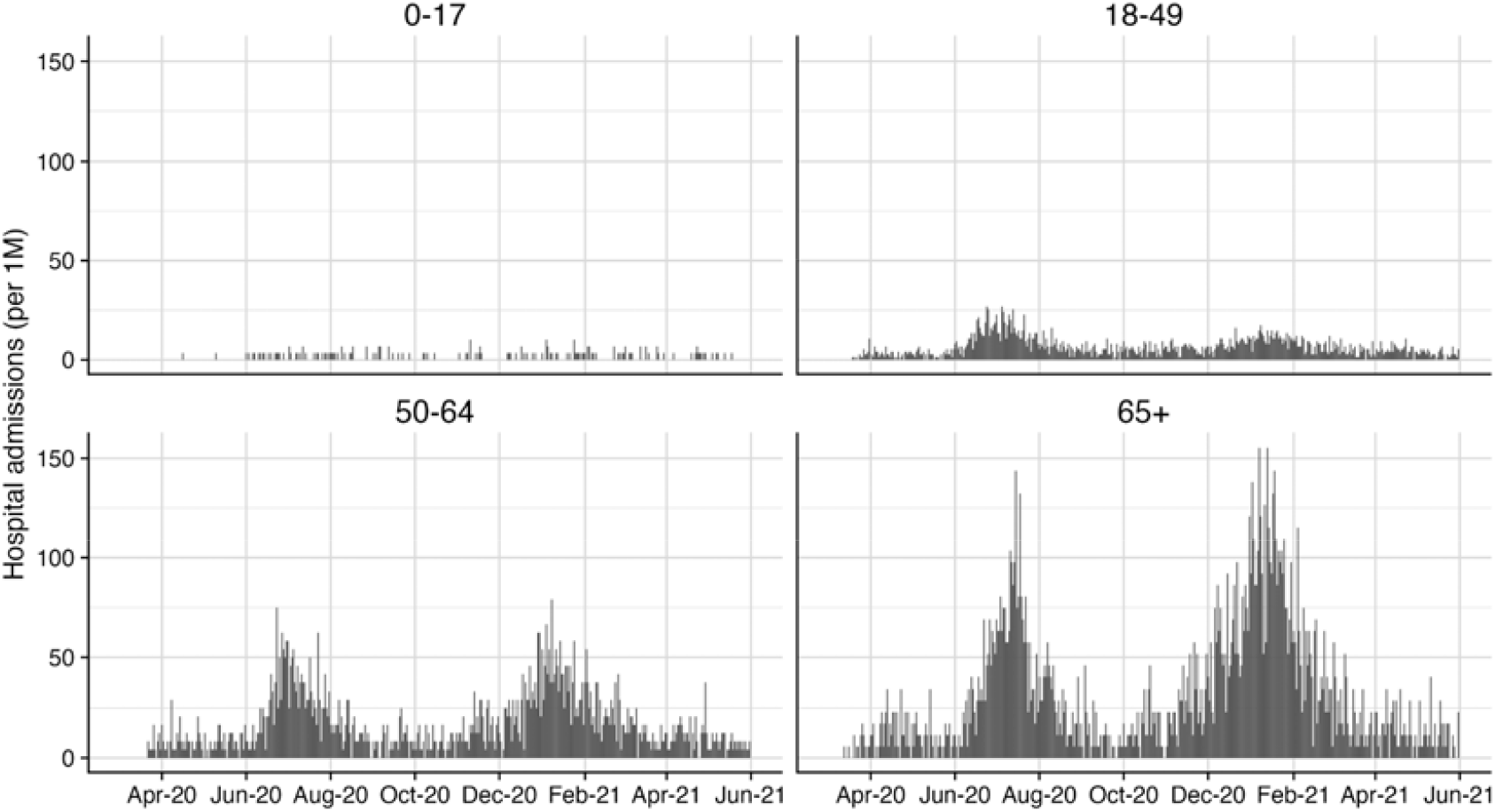
Reported hospital admissions by age group from March 1, 2020 until June 1, 2021. Age-specific admission data provided by Austin Public Health.

**Figure S6:**
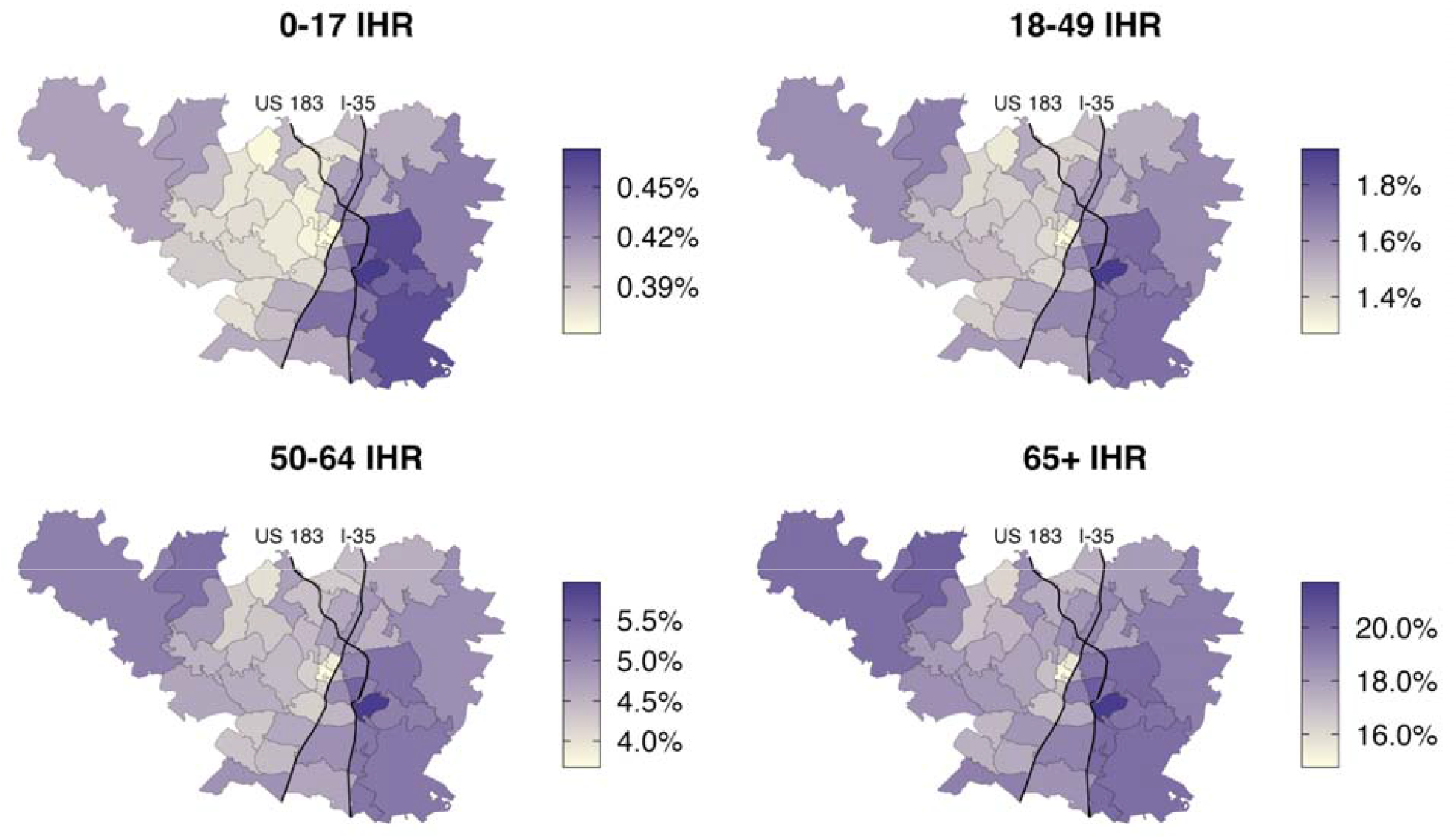
Estimated ZIP code and age-specific IHR for each ZIP code in Travis County. Infection hospitalization rates derived from Texas-specific estimates (Table 1) using population risk estimation methodology for each age group as detailed in [5,54,55].

**Figure S7:**
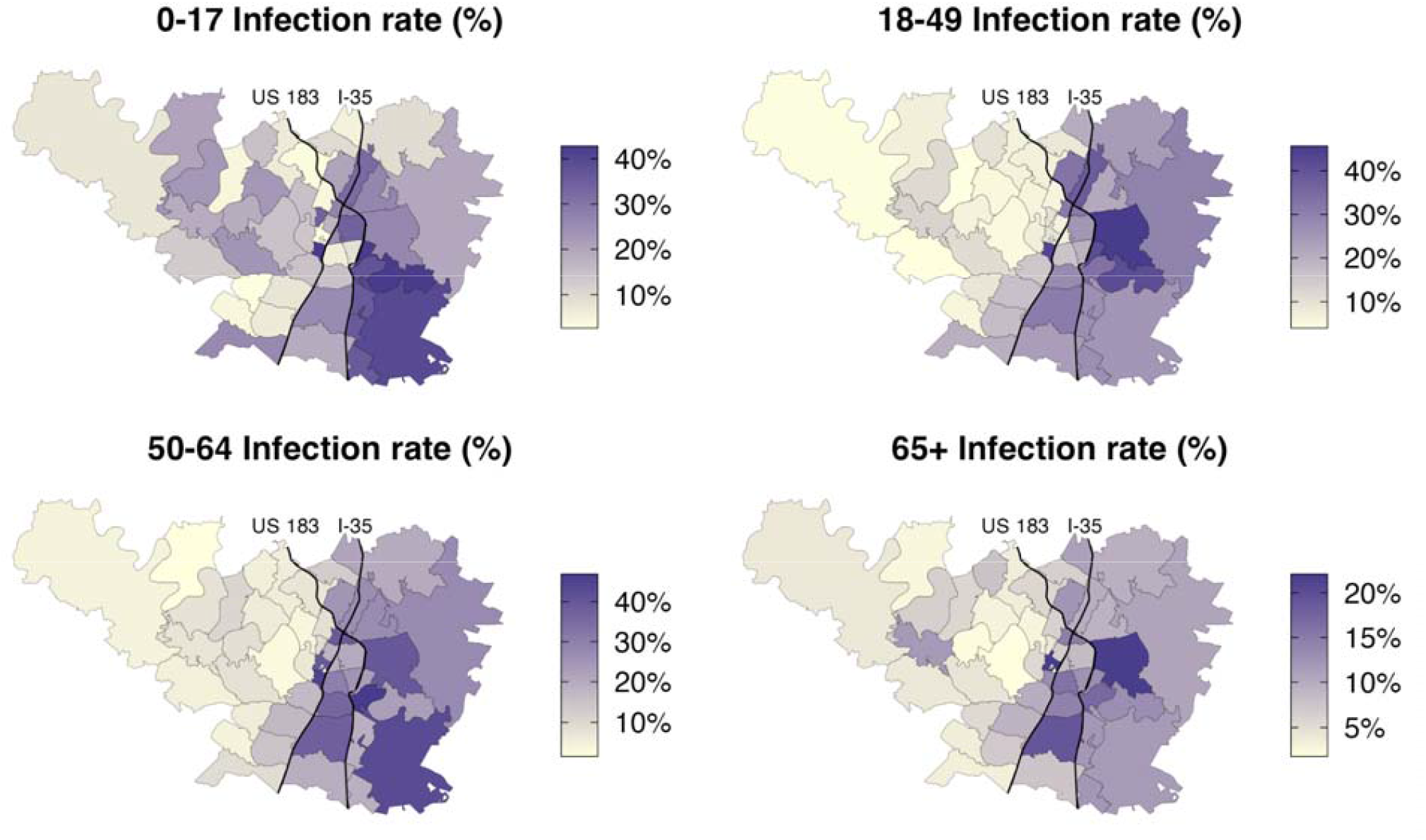
Cumulative infection estimates for each ZIP code and age group in Travis County using hospitalization data up to June 1, 2021.

**Figure S8:**
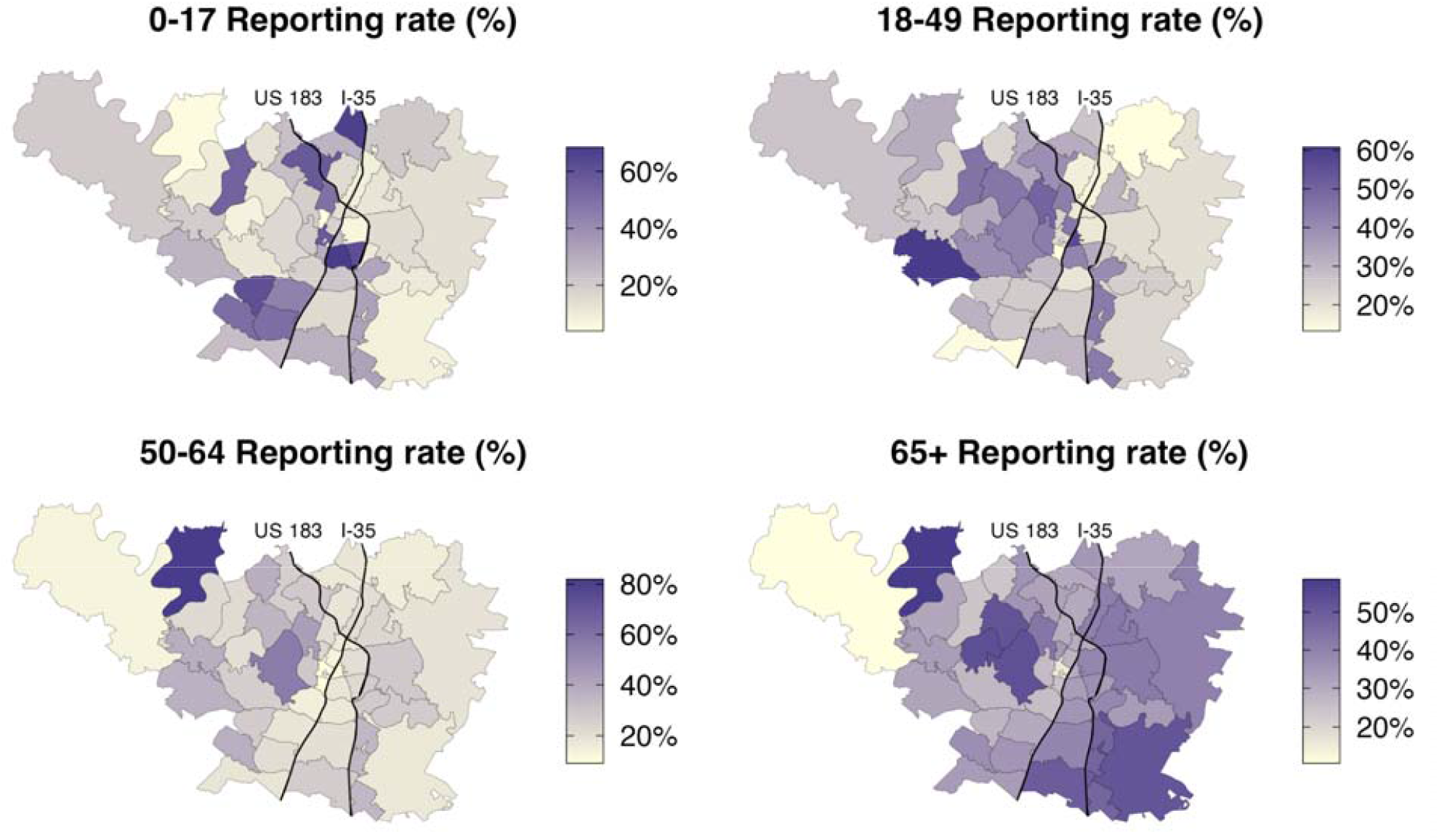
Cumulative estimated reporting rate for each ZIP code and age group in Travis County using reported case data up to June 1, 2021. Testing data used for reporting rates are only a subset of all tests performed, as age and ZIP code stratified data were only available for Austin Public Health administered tests.

**Figure S9:**
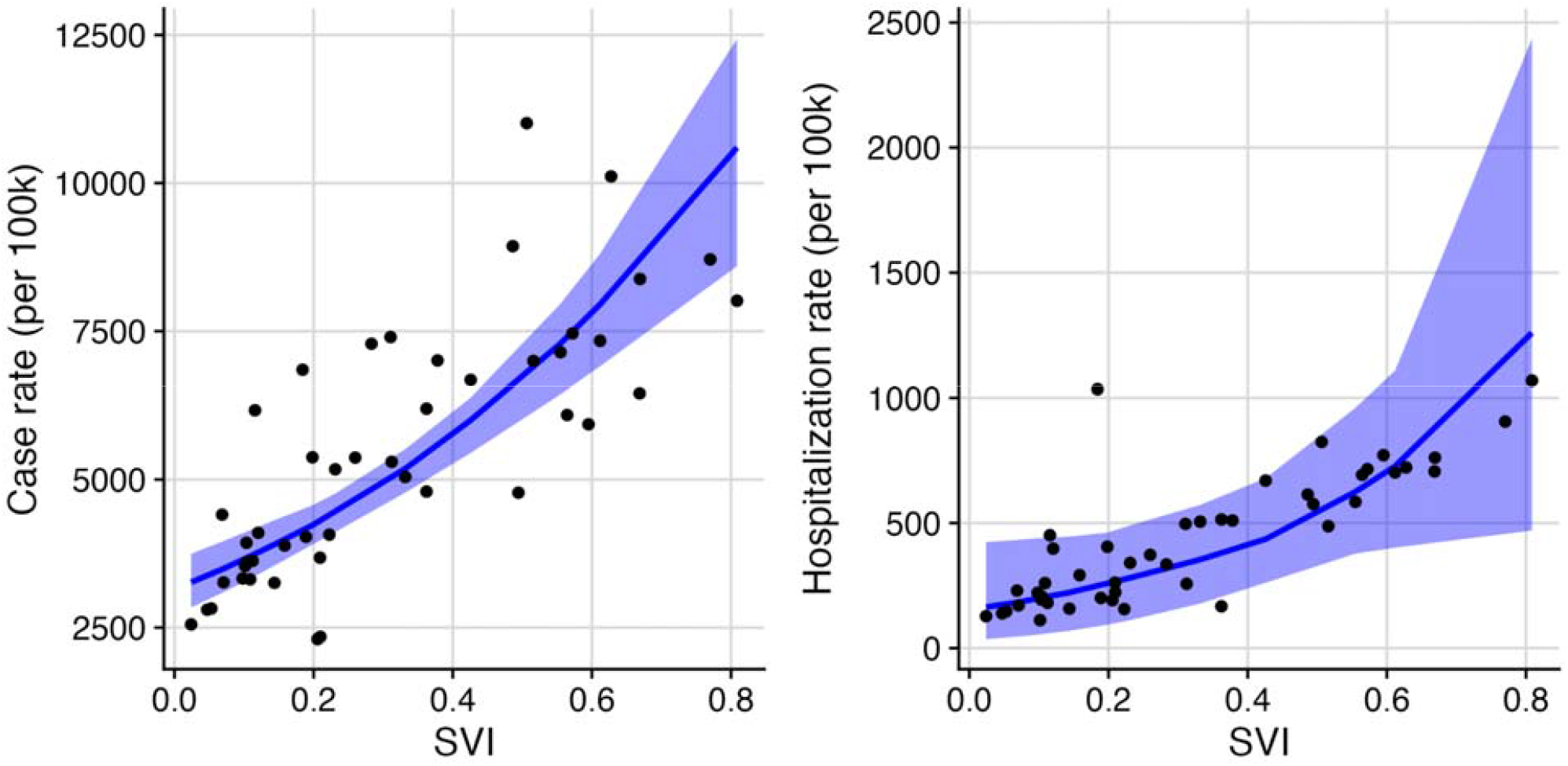
Reported case and hospitalization counts correlate with social vulnerability in Travis County from March 1, 2020 to June 1, 2021. (A) Across the 46 ZIP codes, SVI is a significant predictor of reported case counts (p<0.001). The blue line and ribbon indicate the mean and 95% prediction interval from the fitted Poisson mixed-effects model. (B) Across the 46 ZIP codes, SVI is a significant predictor of reported hospitalization counts (p<0.001). The blue line and ribbon indicate the mean and 95% prediction interval from the fitted Poisson mixed-effects model.

**Figure S10:**
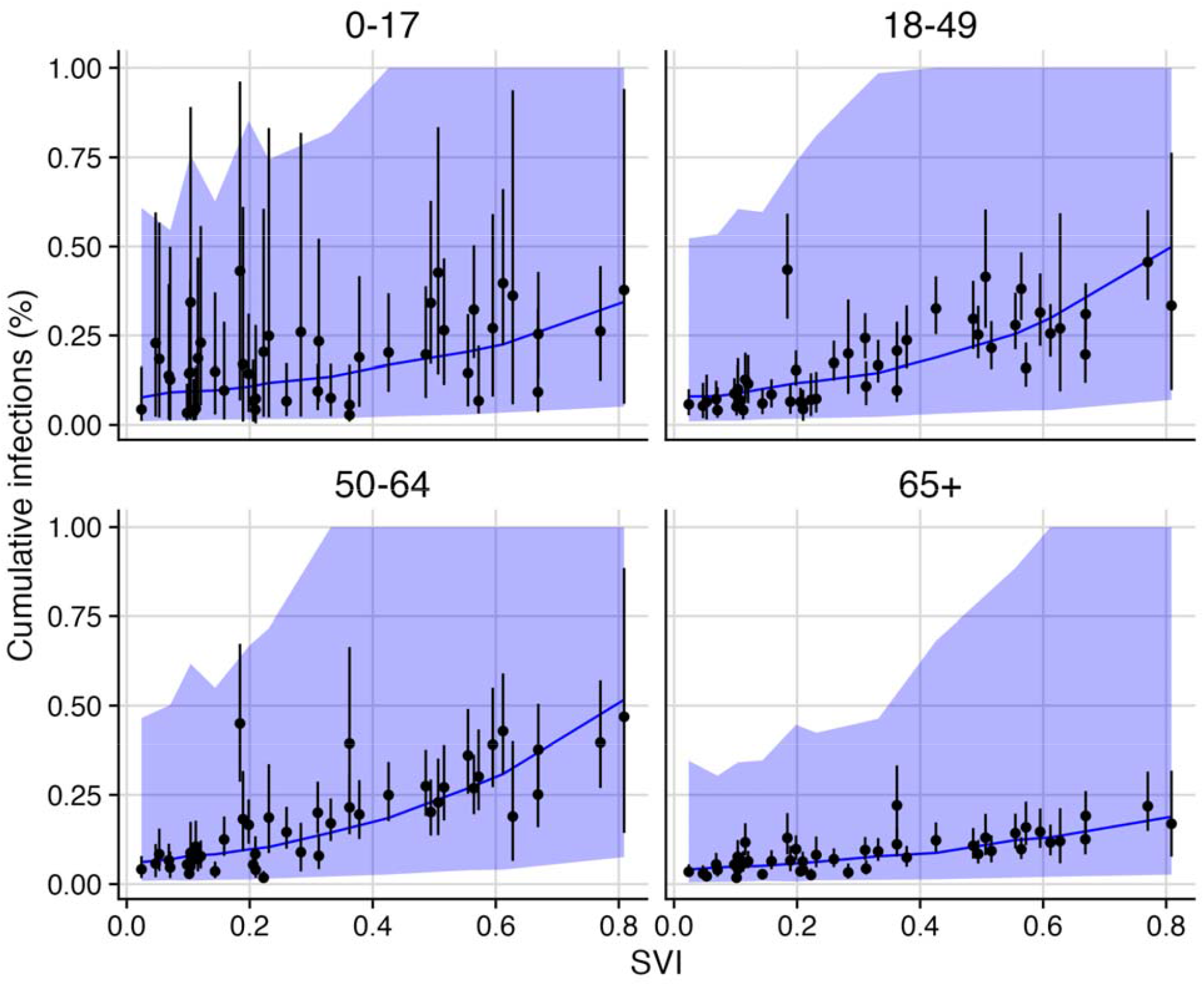
Estimated infection rates correlate with social vulnerability in Travis County from March 1, 2020 to June 1, 2021 across all age groups. Across the 46 ZIP codes, SVI is a significant predictor of reported case counts for every age group (p<0.05, Table S2). Estimated age-specific SVI relationships from the poisson mixed effects regression model are shown in the blue line (mean) and blue ribbon (95% confidence interval).

**Table S2:** Comparison of age-stratified risk ratio between more vulnerable (75th SVI percentile) and less vulnerable (25th SVI percentile) ZIP codes for Travis county, from March 1, 2020 to June 1, 2021. Estimates based on reported COVID-19 hospitalizations are consistently higher than those based on model-derived estimates of ZIP-code level infection rates and observed COVID-19 case rates.

|  | Source of estimate |  |  |  |
| --- | --- | --- | --- | --- |
|  | Estimated COVID-19 reporting rates | Observed COVID-19 case rates | Estimated COVID-19 infection rates | Observed COVID-19 hospitalizations |
| 0-17y | 0.98 (0.90-1.036) | 1.9 (1.6-2.2)* | 2.0 (1.9-2.1)* | 3.8 (2.6-5.5)* |
| 18-49y | 0.88 (0.81-0.95)* | 2.3 (1.9-2.7)* | 2.5 (2.45-2.6)* | 3.2 (1.9-5.2)* |
| 50-64y | 1.02 (0.94-1.11) | 3.1 (2.6-3.6)* | 2.95 (2.9-3.0)* | 3.7 (2.2-6.0)* |
| 65y+ | 1.26 (1.17-1.37)* | 2.8 (2.3-3.3)* | 2.11 (2.0-2.2)* | 2.5 (1.5-3.9)* |

\* Statistically significant (p<0.05)

**Figure S11:**
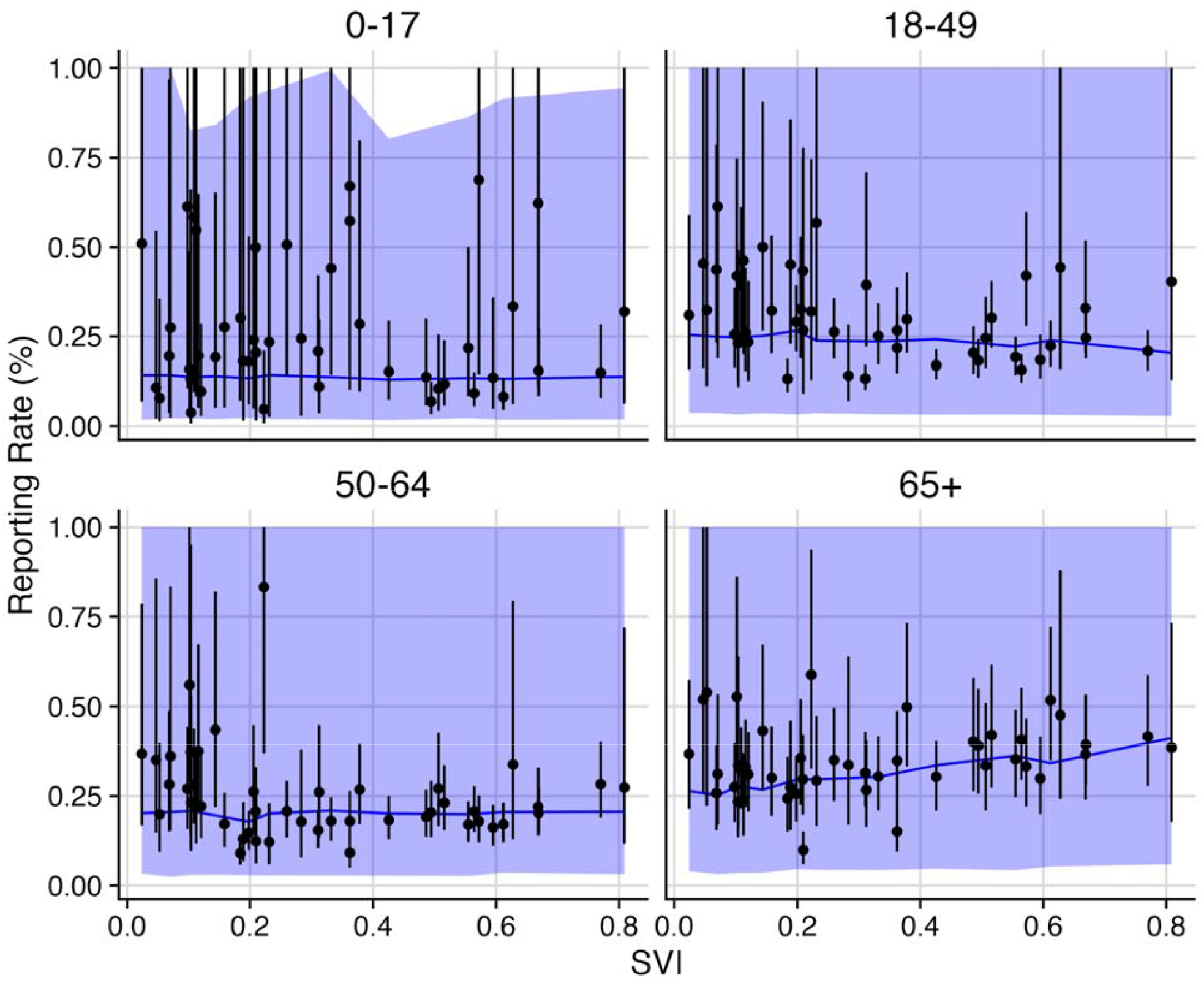
Estimated reporting rates correlate with social vulnerability in Travis County from March 1, 2020 to June 1, 2021. Across the 46 ZIP codes, SVI is a significant predictor of infection reporting rates for the 18-49 and 65+ age groups, with negative relationships estimated for all but the 65+ age group (Table S2). The blue line and ribbon indicate the mean and 95% prediction interval from the fitted Poisson mixed-effects model.

**Figure S12:**
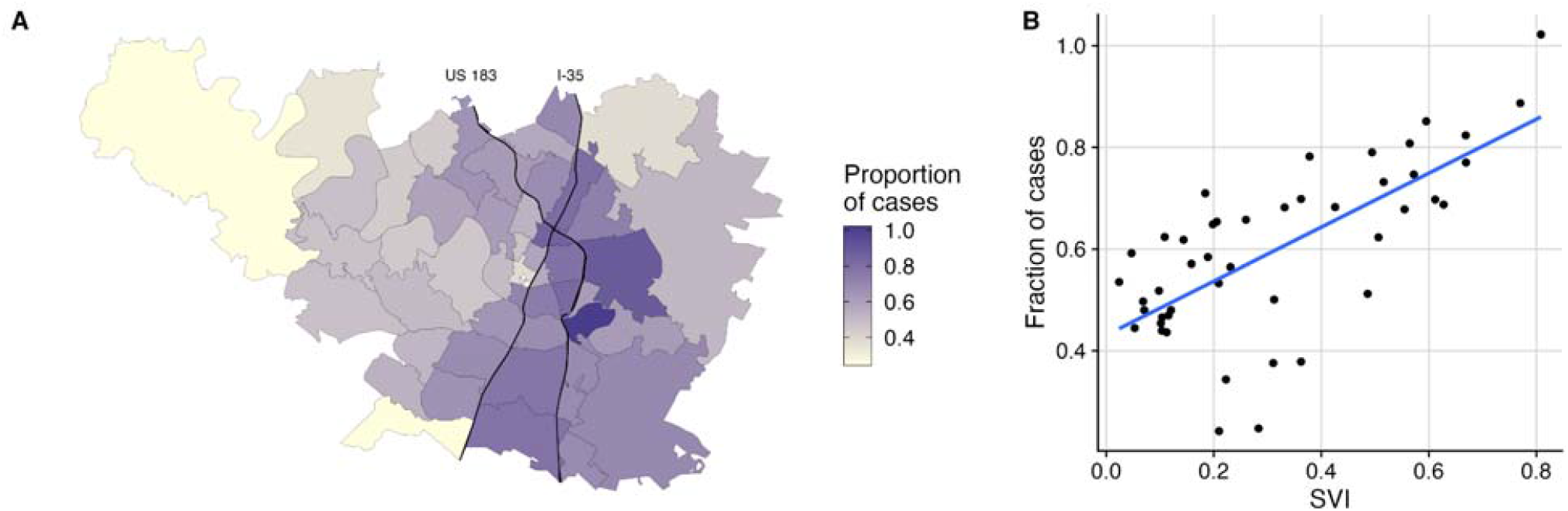
Observed biases in the subset of reported case data stratified by age and ZIP code. (A) Fraction of all reported cases included in the subset of age- and ZIP-code stratified data collected through Austin Public Health’s community testing programs by ZIP code. Overall, the data set covers 60% of all reported cases, but the data set, which does not include all cases identified by private testing sites, has high levels of coverage in the vulnerable ZIP codes of East Austin. (B) Reported case coverage from the dataset correlates positively with SVI. Blue line indicates the mean of a fitted linear regression model.

